# Regular deworming and seasonality are potential challenges but also offer opportunities for hookworm elimination

**DOI:** 10.1101/2023.03.09.23287064

**Authors:** Amanda NL Lamptey, Irene A Larbi, Irene Ofei Owusu Donkor, Jeffrey G Sumboh, Yvonne Ashong, Dickson Osabutey, Michael Cappello, Dennis Adu-Gyasi, Dennis Gyasi Konadu, Kwaku Poku Asante, Kwadwo A Koram, Michael D Wilson

## Abstract

The global health community has targeted the elimination of neglected tropical diseases (NTDs) including soil-transmitted helminthiasis by 2030. The elimination strategy has not changed from that of control using regular mass drug administration (MDA) with albendazole, WASH and education. Already doubts have been expressed about this achievement, principally because drugs do not interrupt transmission. We report here the findings of a cohort study aimed to identify host modifiable and environmental factors associated with hookworm infection and reinfection in rural communities in Kintampo North Municipality, Ghana. Faecal samples of 564 consented participants were screened for intestinal parasites at baseline, 9 months and 24 months using the Kato-Katz method. At each time point, positive cases were treated with a single dose of albendazole (400 mg) and their samples were again screened 10-14 days post-treatment to record treatment failures. The hookworm prevalence at the three-time points was 16.7%, 9.22% and 5.3% respectively, whilst treatment failure rates were 17.25%, 29.03% and 40.9% respectively. The intensities of hookworm infection (in eggs per gram) at the time points were 138.3, 40.5 and 135, which showed a likely association with wet and dry seasons. We posit that the very low intensity of hookworm infections in humans during the dry season offers a window of opportunity for any intervention that could drastically reduce the community worm burden before the rainy season.

## Introduction

Hookworm disease is an important intestinal parasite infection that is highly endemic and ranks the highest in Ghana. [1, 2, 3]. Humans get infected with hookworm when the soil-living third-stage larvae (L3) enter trans-dermally or when they are ingested. Upon entering a human host, L3 larvae migrate through the bloodstream to the lungs, burrow into the alveolar spaces, and are coughed up in mucous and swallowed into the intestinal tract of the host where they eventually mature into adults in the small intestine. The adult hookworm attaches to the host intestinal mucosa and using its cutting apparatus disrupts capillaries both mechanically and enzymatically while sustaining blood flow with the use of anticoagulant substances [4]. Adult male and female worms mate in the intestines and the female worm may produce up to 20,000 eggs per day, which are passed into the host’s faeces [5]. The clinical symptoms are iron deficiency anaemia due to blood loss, abdominal pain, diarrhoea, and protein malnutrition, which lead to impaired physical and cognitive development in children, and increased perinatal maternal/infant mortalities in pregnant women [6].

Current control efforts recommended by the WHO for hookworm and other STHs are regular deworming with single doses of albendazole (400 mg) or of mebendazole (500 mg) in preventative chemotherapy campaigns [7]. This population-based MDA with benzimidazoles for STH has proven effective in the treatment of human hookworm infection but in recent times there have been reports of reduced efficacy of albendazole in the treatment of soil-transmitted helminths [8].

Hookworms are among the group of STH and other NTDs that are earmarked for elimination by the year 2030, which Ghana has also endorsed. There are already doubts that this may not be achieved for multiple reasons, the principal among these is that MDA does not interrupt transmission, as reviewed by Haldemann et al. [9].

We report here the findings of a cohort study that was aimed at investigating host-modifiable factors associated with hookworm infection and reinfection, and albendazole treatment response in nine rural communities in the Kintampo North Municipality in the Bono East Region of Ghana. We found that albendazole treatment whilst reducing the prevalence also leads to increased non-responsiveness to the drug and that the intensity of infections in humans is very low in the dry and high in the wet season.

## Materials and Methods

### Ethical statement

A formal informed consent was obtained from all the participants and in the case of children, both assent and parent/guardian approvals were sought. The information was given verbally in the local language and their approvals were affirmed by fingerprinting in the case of illiterate participants.The study received approvals from the Noguchi Memorial Institute for Medical Research Ethics Review Board (NIRB #100/16-17), the Kintampo Health Research Center Ethics Review Committee (# KHRCIEC 2017-20) and NIH/NIAID (DMID Protocol# 17-0061).

### Study sites and population

The study was conducted in nine rural communities in the Kintampo North Municipality of the Bono Region of Ghana (Fig 1).

**Fig 1.**
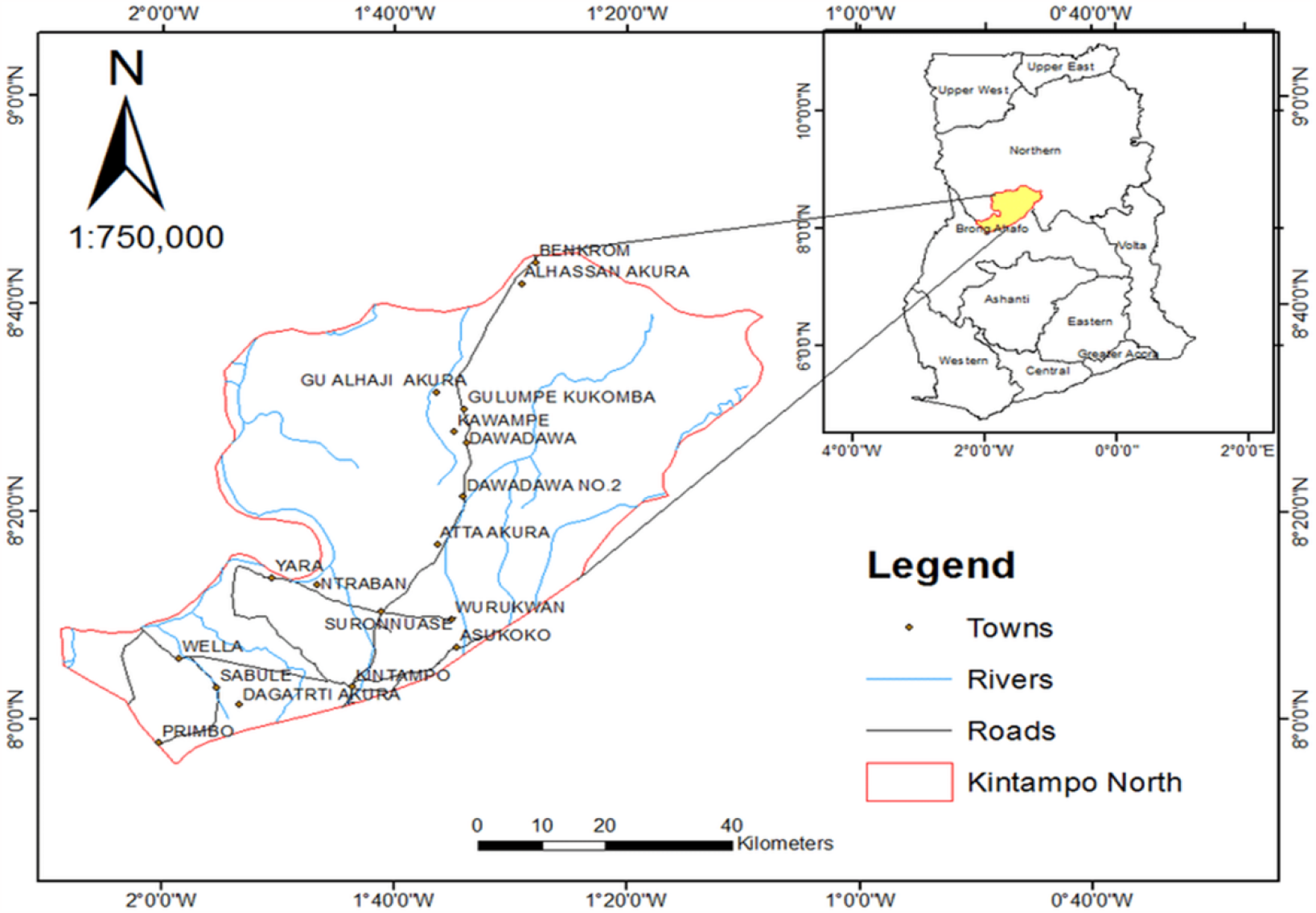
Map of the Kintampo North and South Municipalities and communities showing their locations in Ghana and where the cohort studies were conducted.

### Study population

Five hundred and sixty consented participants comprising 279 (49.82%) males and 281 (50.18%) females aged between 5.59 - 93.15 years were initially recruited into the cohort study.

### Faecal examination and analysis

The faecal sample of each study participant who presented was collected and examined for intestinal parasites using the Kato-Katz faecal thick smear technique [10]. Duplicate slides were prepared for all samples, which were read within 30–60 minutes by two experienced microscopists. A third reading was made if the scores of the two microscopists differed significantly otherwise the mean value was recorded. Hookworm-positive individuals were treated with single-dose albendazole and their stool specimens were collected 10–14 days post-treatment and examined using the Kato-Katz method as described above. The same process was repeated at baseline, 9 months and 24-month time points. The prevalence was determined from the number of positives and the intensity of infections expressed as the number of eggs per gram (EPG) of faeces was also determined from the counts of only positive cases (i.e. negative cases were excluded), and so only the arithmetic means were calculated and reported in this study. The treatment failure rate [11] was calculated from the number of infected persons out of the total treated at each time point

## Results

Of the 564 participants who gave faecal samples 94 were infected at baseline and the intensities of infection were 89 (93.68%) light; 4 (4.21%) moderate and 2 (2.11%) heavy 89 (93.68%). The recorded hookworm prevalence at the three-time points was 16.7% (95/564), 9.22% (48/524) and 5.3% (26/495) respectively and the corresponding albendazole treatment failure rates were 17.25% (14/78), 29.03% (9/31) and 40.9% (9/22) respectively (Fig 2).

**Fig 2.**
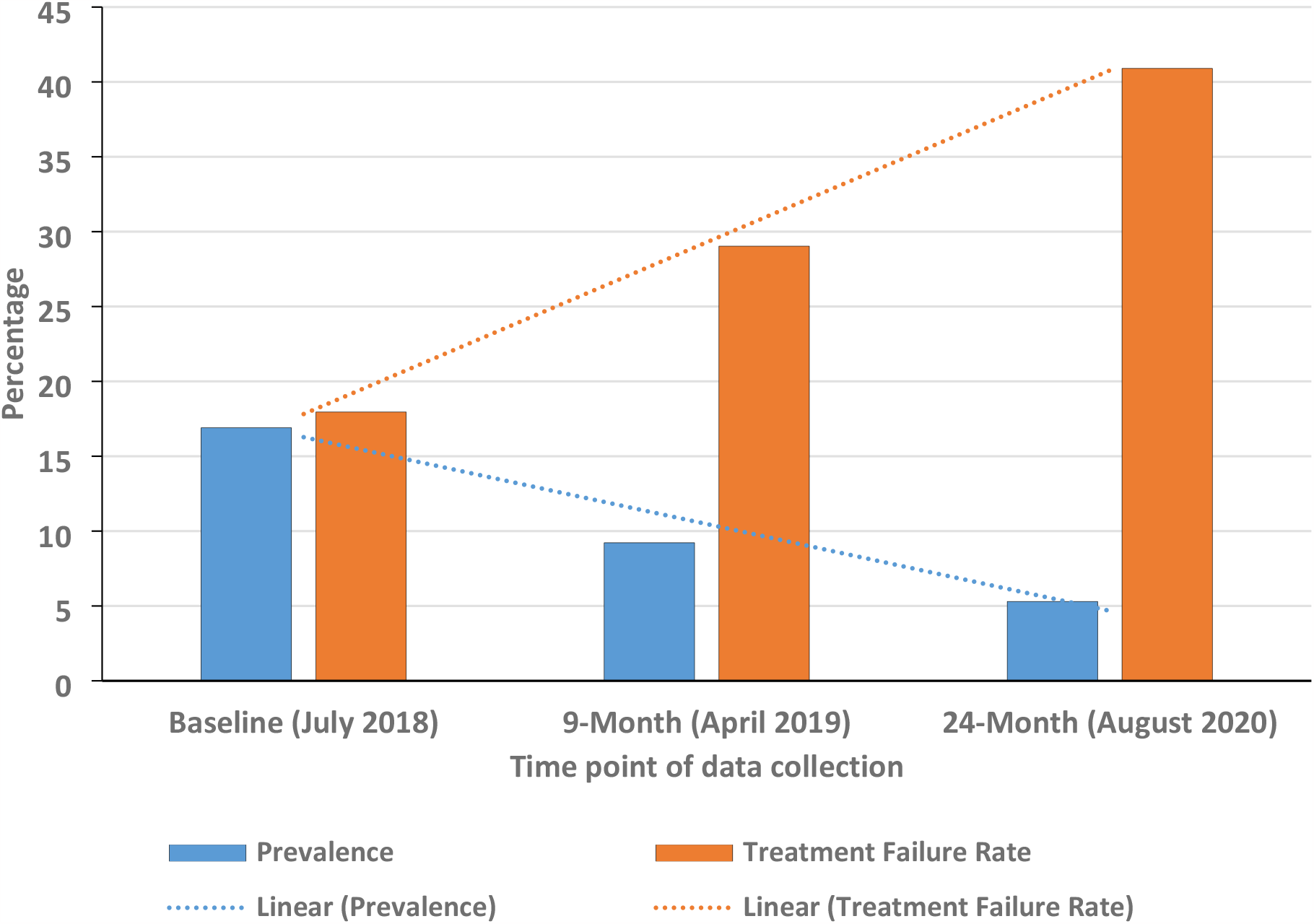
Graph showing the trends in the hookworm prevalence and the albendazole treatment failures across all the three-time points. The prevalence of hookworm infections fell by 68.9% whilst that of treatment failures increased by 1.37-fold.

The intensities of infection were 138.3 e.p.g., 40.5 e.p.g. and 135 e.p.g. respectively at the time points (Figure 3). Figure 3 shows a significant reduction in the intensity of infections during the 9-month time point compared to baseline and the 24-month time points, both of which were almost at the same level.

**Fig 3.**
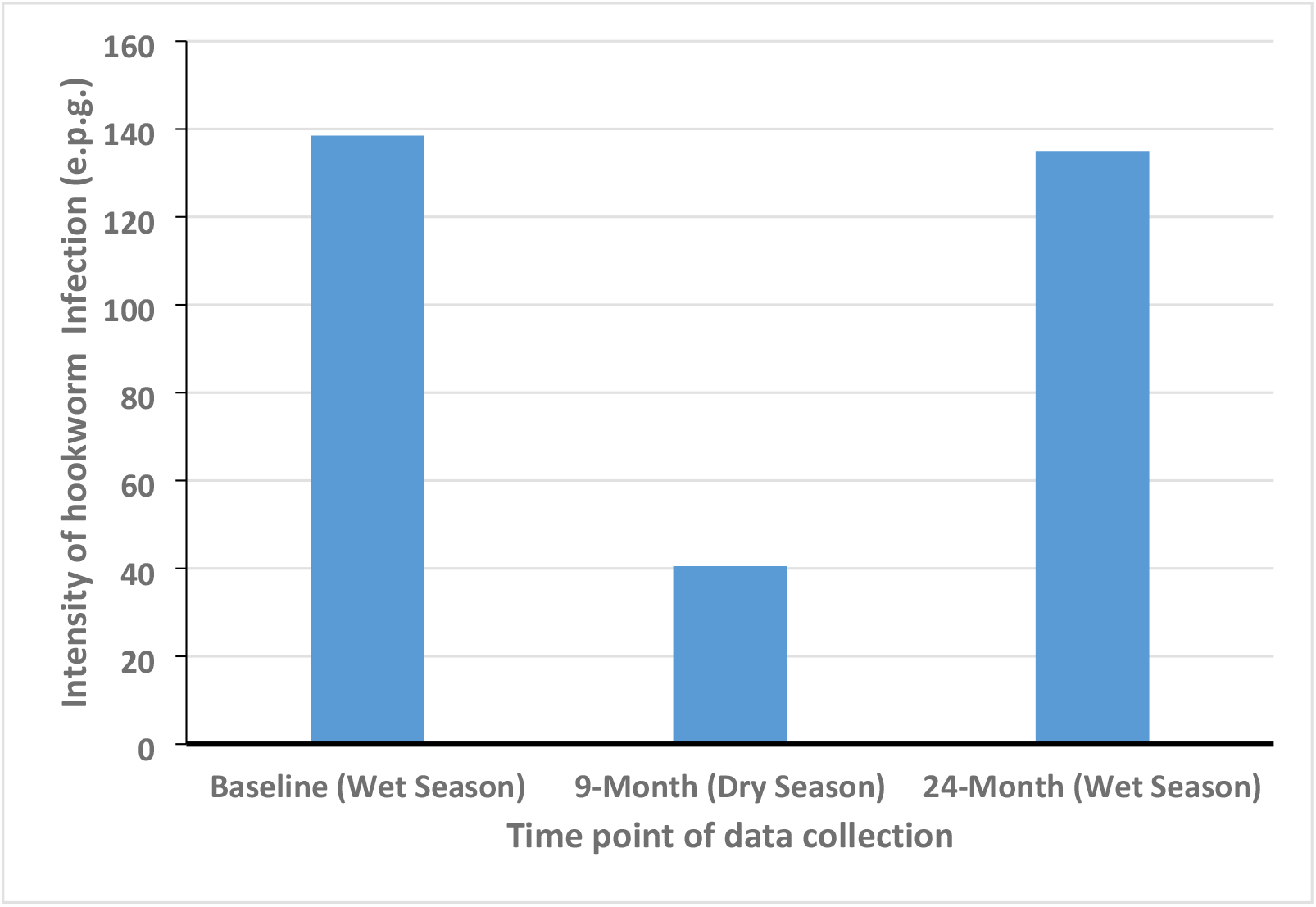
Graph showing the trend in the mean hookworm intensities across all three-time points. It shows a reduction in the intensity of hookworm infections in humans in the dry season.

## Discussion

Despite the control efforts initiated decades ago against STH in endemic countries we are yet to have evidence of any successful elimination using MDA. In attempts to drive down STH prevalence, WASH and health education were added measures to MDA that were expanded to include all school-age individuals in endemic communities. Our previous studies and those of others have shown that MDA with benzimidazoles alone cannot drive the transmission of STH to zero. There is, therefore, the need to identify factors associated with the transmission, which when targeted would further drive down STH prevalence.

Rather worrying for the global health community is our finding that whilst deworming reduces hookworm prevalence, the rates of non-responsiveness increase and if by extension this is what regular MDA does in the field. We have shown in our previous publication that at a high concentration of drug (5 mg/mL) the median hookworm eggs hatch rate was 2.3 fold lower in post-treatment samples when compared with those collected before treatment (14.8% versus 34.0%) [2], which corroborates the observed effect of albendazole of the present study. One potential mechanism for this observation is the possibility that albendazole treatment selects for drug resistance in hookworms, which could explain why the “treatment failure” rate increased with each subsequent time point. It, therefore, poses a challenge to the elimination strategy of using MDA in the long term.

By a fortunate stroke of serendipity, the 9-month data collection occurred during the dry season whilst those of the baseline and the 24-month were during the wet season. The observed dip in the hookworm intensities in the dry season suggests that the community burden of the parasite is low during this time. Interestingly, the recorded intensities of infections were similar during the wet season at baseline and the 24-month time point which is also suggestive of an intense transmission during the rainy season. This finding is corroborated by a study conducted in the Gambia, which observed a steady rise in the intensity of hookworm infection after the onset of the rains [12]. The reason for the increase in the fecundity of adult hookworms during the rainy season is unknown but it might be due to any of the two possible mechanisms that have been observed in the veterinary field. Shaw and Moss observed seasonal variations in worm fecundity but concluded that the high outputs during spring/summer may be due to increased transmission leading to additional infections with fresh threadworms (*Trichostrongylus tenuis*) in red grouse [13], and also corroborated by Ogbourne that the level of *Trichonema catinatum* among horses is maintained by the arrival of fresh individuals during the summer [14]. This phenomenon is supported by the observation that changes in the size of parasitic populations of *Trichonema nassatum* follow seasonal variations in the rate of infection, as more individual worms mature during summer/autumn than during winter/spring because of proportional differences in the numbers of infective larvae ingested from the pasture [15]. The other possible phenomenon is suggestive of diapause, whereby *T. longibursutum, T. catinatum* and *T. goldi* larvae ingested by grazing horses during summer accumulate in the gut wall with their development arrested until the following spring when 4th∼stage larvae emerge collectively and quickly mature to adults [15].

Irrespective of the mechanism at play in hookworm transmission, the opportunity that this offers for hookworm elimination is to implement efforts to drastically reduce the parasite load in communities before the onset of the rainy season and that includes mass deworming in endemic communities.

## Data Availability

All data underlying the findings reported in the submitted manuscript is provided as part of the submitted article.

## Acknowledgements

We appreciate the support of the NIINE laboratory and fieldwork teams, along with that provided by the staff of the Kintampo Health and Research Centre. We are also grateful to the chiefs, community leaders and participants in the Kintampo-North Municipality.

## Competing interests

The authors declare no competing interests.

## Funding

The study was funded by NIH/NIAID awards to MDW (U19AI129916) and MC (R01AI132452). The funding agency did not play any role in the design and execution of the study.

## Authors’ Contributions

MDW and MC conceived the project. JGS, DO, YA conducted the field and laboratory works, DA-G, DGK, IAL and IOOD performed the data analysis, AL drafted the manuscript and all the authors read and approved it.

## Notes

### Competing Interest Statement

The authors have declared no competing interest.

### Author Declarations

The study received approvals from the Noguchi Memorial Institute for Medical Research Ethics Review Board (NIRB #100/16-17), the Kintampo Health Research Center Ethics Review Committee (# KHRCIEC 2017-20) and NIH/NIAID (DMID Protocol# 17-0061).

